# Changes in self-reported alcohol consumption at high and low consumption in the wake of the COVID-19 pandemic: A test of the polarization hypothesis

**DOI:** 10.1101/2024.07.31.24311291

**Authors:** Alexander Tran, Huan Jiang, Shannon Lange, Mindaugas Štelemėkas, Daumantas Stumbrys, Ilona Tamutienė, Jürgen Rehm

## Abstract

**Background:** The Coronavirus Disease 2019 (COVID-19) pandemic and associated public health measures had an impact on alcohol use. Based on the literature of past crises (health, economic, etc.), it was hypothesized that the COVID-19 pandemic led to a polarization of drinking–that is, heavy drinkers increased their drinking, while light to moderate drinkers decreased their drinking and/or temporarily abstained. The aim of the current study was to test the respective hypothesis.

**Methods:** Data from the Reducing Alcohol Related Harm Standard European Alcohol Survey for Lithuania were obtained for 2015 and 2020. Average daily consumption (in grams per day) was decomposed into deciles for each year, and compared pre-COVID to onset-of-COVID pandemic across the highest, second highest, and lowest deciles. A comparison of population-levels of mental health was conducted between pre-COVID and the onset-of-COVID.

**Results:** The findings indicated that overall, there was higher consumption in 2015, M_2015_ = 11.49 (SD = 8.23) vs. M_2020_ = 10.71 (SD = 12.12), p < .00001. However the opposite was found in the highest decile M_2015_ = 29.26 (SD = 5.44) vs. M_2020_ = 39.23 (SD = 20.58), *p* = .0003. This reversal pattern was not observed in the second highest nor the lowest decile. There was a lower proportion of respondents indicating “bad” mental health pre- vs.post-COVID (3.4% vs. 6.5%).

**Conclusion:** Although COVID was associated with nationwide declines in alcohol consumption, this was not the case for all segments of the population. In Lithuania, it appears that there was an increase in consumption among the heaviest drinkers, supporting the polarization hypothesis.

## Introduction

During the Coronavirus Disease 2019 (COVID-19) pandemic there was a considerable shift in behavior as people experienced fear, anxiety, and uncertainty around this novel disease (Coelho et al., 2020; Erbiçer et al., 2022; McBride et al., 2021). Across the globe, countries implemented lockdowns to prevent the spread of the disease which, among many consequences, led to widespread social isolation (Ganesan et al., 2021; Onyeaka et al., 2021; Verma et al., 2020). An important question regarding the outcomes of the lockdown is how peoples’ health behavior, such as alcohol use, may have been impacted (McBride, et al., 2021).

COVID-19 associated lockdowns and other measures, had a complex effect on alcohol consumption and alcohol-attributable harm. In the beginning of the epidemic, in March 2020, it had been predicted (Rehm et al., 2020) that the effects would be primarily twofold, overall, there would be less availability and affordability of alcohol, leading to a decrease in consumption (for details on these mechanisms see (Babor et al., 2022)). On the other hand, in previous economic and natural crisis situations, alcohol was used as coping mechanism for the stress, fear and anxiety as would be the case with a novel pandemic like COVID-19 (see (De Goeij et al., 2015; Pohorecky, 1991) for alcohol as coping mechanism). Together, both mechanisms were predicted to lead to polarization of drinking, meaning that overall we see a reduction of alcohol consumption, but certain groups, especially groups with prior heavier drinking patterns and/or mental problems, would increase their consumption (Rehm, 2024).

In the first year of COVID-19 alone, global consumption indeed went down by 10.3% and in the EU by 6.6% (World Health Organization, 2024). However researchers found that while some of the population dramatically decreased their consumption, for others, there was an increase (Bloomfield et al., 2022; Kilian et al., 2022; Kilian et al., 2021; Neufeld et al., 2020; Sohi et al., 2022). Thus, although in general consumption may have shown a decline or no change following the onset of COVID-19, analyzing the trends within sub populations could improve the understanding of how specific individuals were affected. For instance, heavy drinkers might have a sudden increase in access to alcohol consumption via home delivery or those prone to coping with stress using alcohol could have experienced heightened mental distress during this period of uncertainty.

The primary aim of the current study was to test the polarization of drinking hypothesis–that is, heavy drinkers increased their drinking, while light to moderate drinkers decreased their drinking and/or temporarily abstained–using data from Lithuania. A secondary aim, was to determine how the self-reported mental health of the general population changed from pre-COVID to the onset-of-COVID.

## Methods

### Data source

Data were collected from the Reducing Alcohol Related Harm Standard European Alcohol Survey (RARHA SEAS). The RARHA SEAS survey was a standardized survey distributed across 33 EU countries that measured respondents drinking patterns, drinking preferences, history with alcohol, motivations for drinking, general health, and demographic information. We analyzed data for Lithuania only. In total there were 2136 responses across both surveys which were split into pre-COVID (data collection year 2015, n=1313) and post-COVID (data collection year, 2020, n=823). For this analysis, we focused on estimated daily self-reported alcohol consumption and self-reported mental health. The survey measures beverage-specific alcohol consumption (beer, wine, spirits) based on frequency and amount and the values were aggregated into a measure of average daily alcohol consumption. Values were capped when computing estimated daily alcohol consumption, with a maximum of 50cl of pure alcohol per beverage type (about 28 standard drinks, see https://www.rarha.eu/NewsEvents/LatestNews/Lists/LatestNews/Attachments/37/RARHA%20SEAS%20Data%20base%20-%20guidelines_rev1.pdf for full description of variables).

### Analysis

To test the hypothesis that COVID led to polarization we first performed a Kolmogorov-Smirnov (KS) test to determine if the distributions of the alcohol consumption responses differed between the 2015 and the 2020 surveys (Lilliefors, 1967). For non-parametric distributions, we followed this analysis up with a Mann-Whitney U (MWU) test (Hettmansperger & McKean, 2010) to compare the average values of daily consumption between the years. Next, we separated daily alcohol consumption into deciles and performed the same test for the highest, second highest, and lowest decile values. For the secondary hypothesis aimed at observing differences in mental health, we measured the proportion of responses for the mental health item across the two surveys. The item asked “How would you rate your psychological well-being?”on a Likert-type scale (1 – Very good, 2- Good, 3- Fair, 4 – Bad, 5 – Very bad, 9 – No answer). Responses were compared between surveys and further analyzed using a Chi-squared 2-sample test for equality of proportions. All analyses were stratified by sex and conducted in R Version 4.1.5 (R Development Core Team, 2023).

## Results

The KS test determined that the two surveys had significantly different distributions (D = .14, p < .00001, see Figure 1). The MWU test showed that there was significantly higher alcohol consumption in 2015 than in 2020 (p < .00001, M_2015_ = 11.49±8.23 vs. M_2020_ = 10.71±12.12 cl of pure alcohol) equating to roughly 6.5 and 6 standard drinks per day (1 standard drink = 14 grams of pure alcohol), respectively.

**Figure 1.**
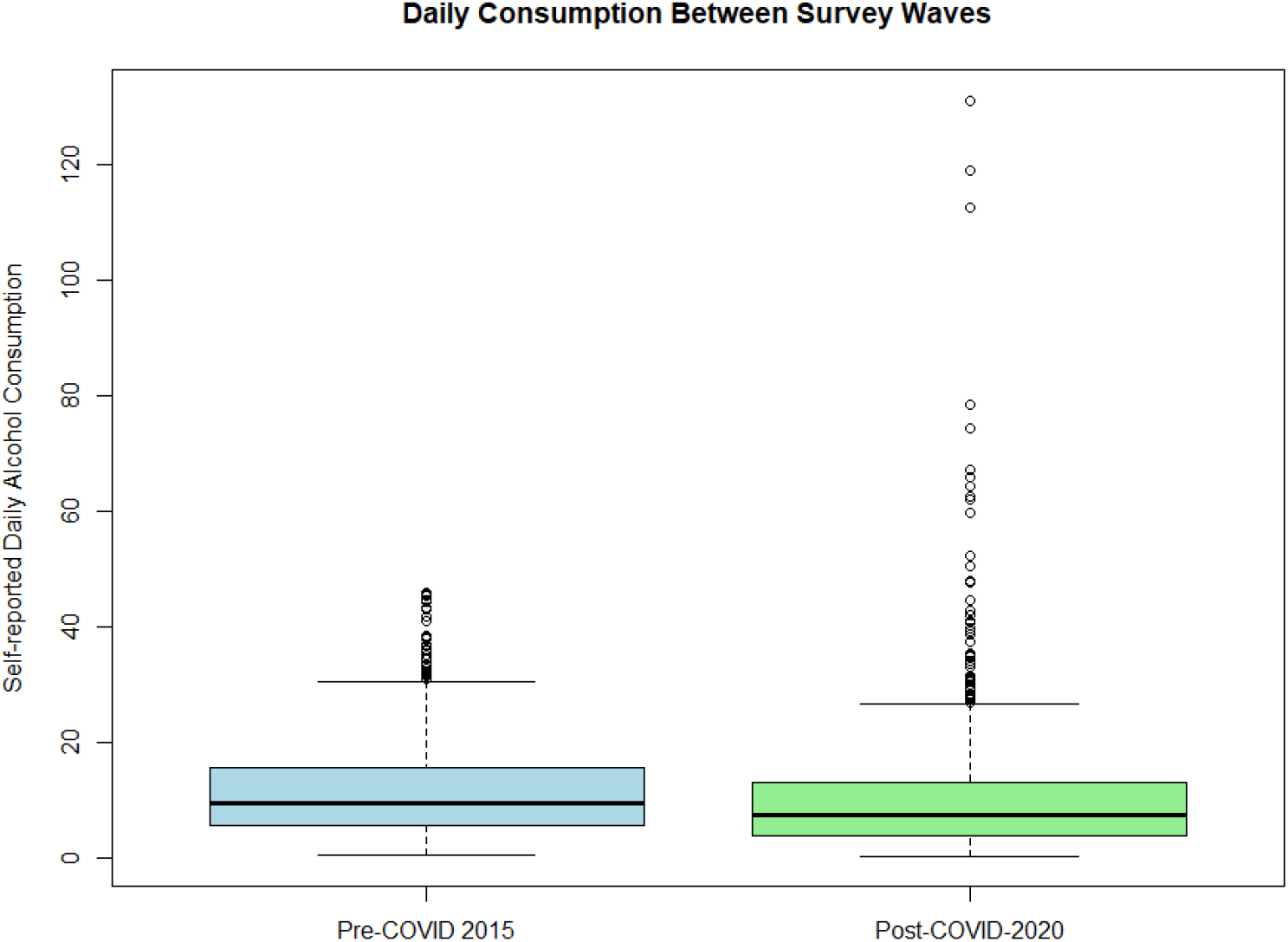
Distribution of values for daily alcohol consumption (in cl) pre and post COVID as a Box-and Whisker plot (Panel A)

The data were then split into deciles of daily alcohol consumption (see Figure 2). To test the polarization hypothesis of whether there was increased drinking at the upper decile during COVID, a KS-test was performed on only those in the 10^th^ decile, followed by a MWU test. The KS-test showed significantly different distributions (D =.30, p = .0002), and the MWU test showed significantly higher consumption during COVID (p = .0003, M_2015_ = 29.26±5.44 vs. M_2020_ = 39.23±20.58 cl of pure alcohol) equating to roughly 16.5 vs. 22 standard drinks per day, compared to before COVID. The tests were repeated for the lowest decile, and the second highest decile (9^th^ decile), which indicated that there was significantly higher alcohol consumption in 2015 (see Table 1), compared to 2020.

**Figure 2.**
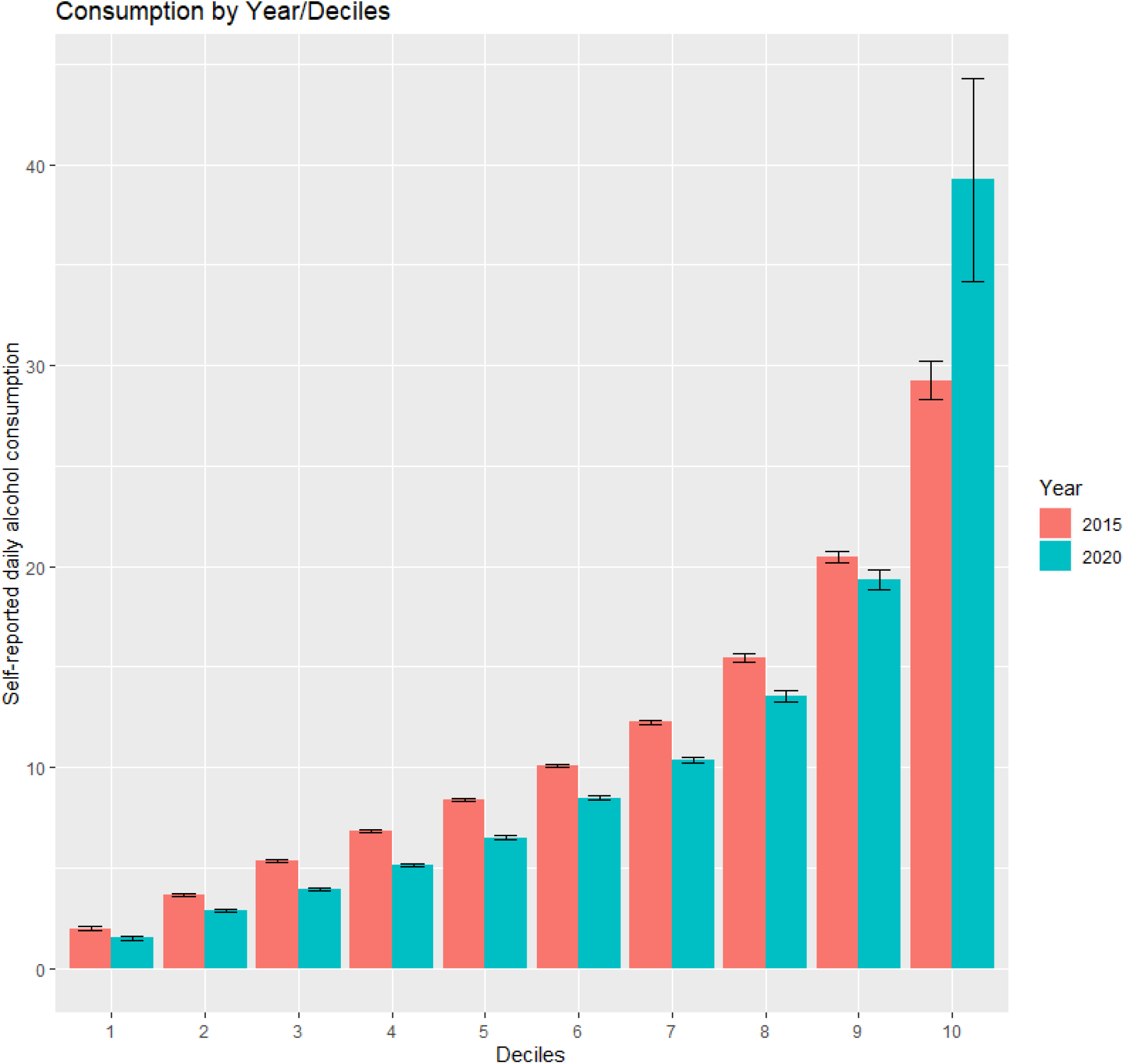
Alcohol consumption separated by deciles compared between pre- and post-COVID (2015 vs. 2020)

**Table 1.** Analysis of change in alcohol consumption pre-and post-COVID, means, and SD of consumption for the 1^st^ and 9^th^ deciles.

|  | Test type | statistic | p-value | Consumption in cl 2015<br>(M±SD) | Consumption in cl 2020<br>(M±SD) |
| --- | --- | --- | --- | --- | --- |
| Decile 1 | KS-test, | D = .41, | $p < .00001$ | 1.98±0.66 | 1.51±0.51 |
| | MWU test | W = 7365 | $p < .00001$ | | |
| Decile 9 | KS-test, | D = .32, | $p < .0001$ | 20.49±1.70 | 19.33±2.25 |
| | MWU test | W = 7299 | $p < .0001$ | | |

Next, the data were separated by sex and the upper decile was analyzed. The results showed that males consumed more alcohol than females in upper decile in 2015 (KS-test, D = .40, p < .0001, MWU-test, p < .0001, M_males_ = 25.49±6.16 vs. M_females_ = 21.40±2.65), however in 2020, there was no significant difference between sexes (KS-test, D = .15, p = .38, t(75) = .24, p = .81, M_males_ = 28.85±18.02 vs. M_females_ = 28.09±17.40, see Figure 3).

**Figure 3.**
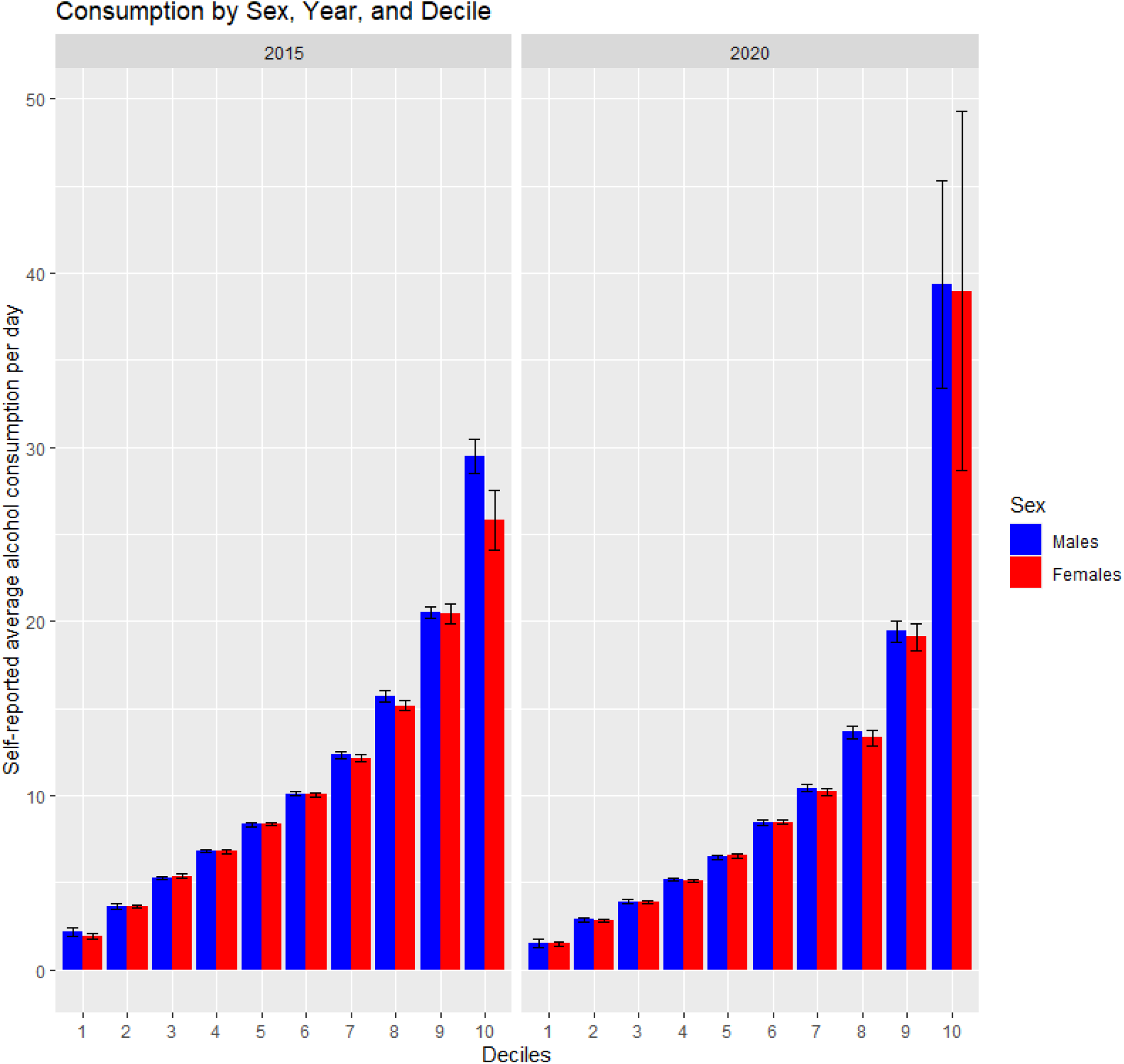
Alcohol consumption separated by deciles and stratified by sex, between pre- and post-COVID (2015 vs. 2020)

Finally, we compared the of responses for a mental health item across the two surveys (see Figure 4). There was a notable increase in respondents that reported “Bad” mental health, and thus we used a proportion test to whether there was a significant increase in proportion of those with bad mental health from 2015 to 2020. The Chi-squared test for equality of proportions indicated there was a significantly high proportion of people reporting bad mental health in 2020 (6.5%) as compared to 2015 (3.4%, Chi-χ^2^ = 10.65, p = .001).

**Figure 4.**
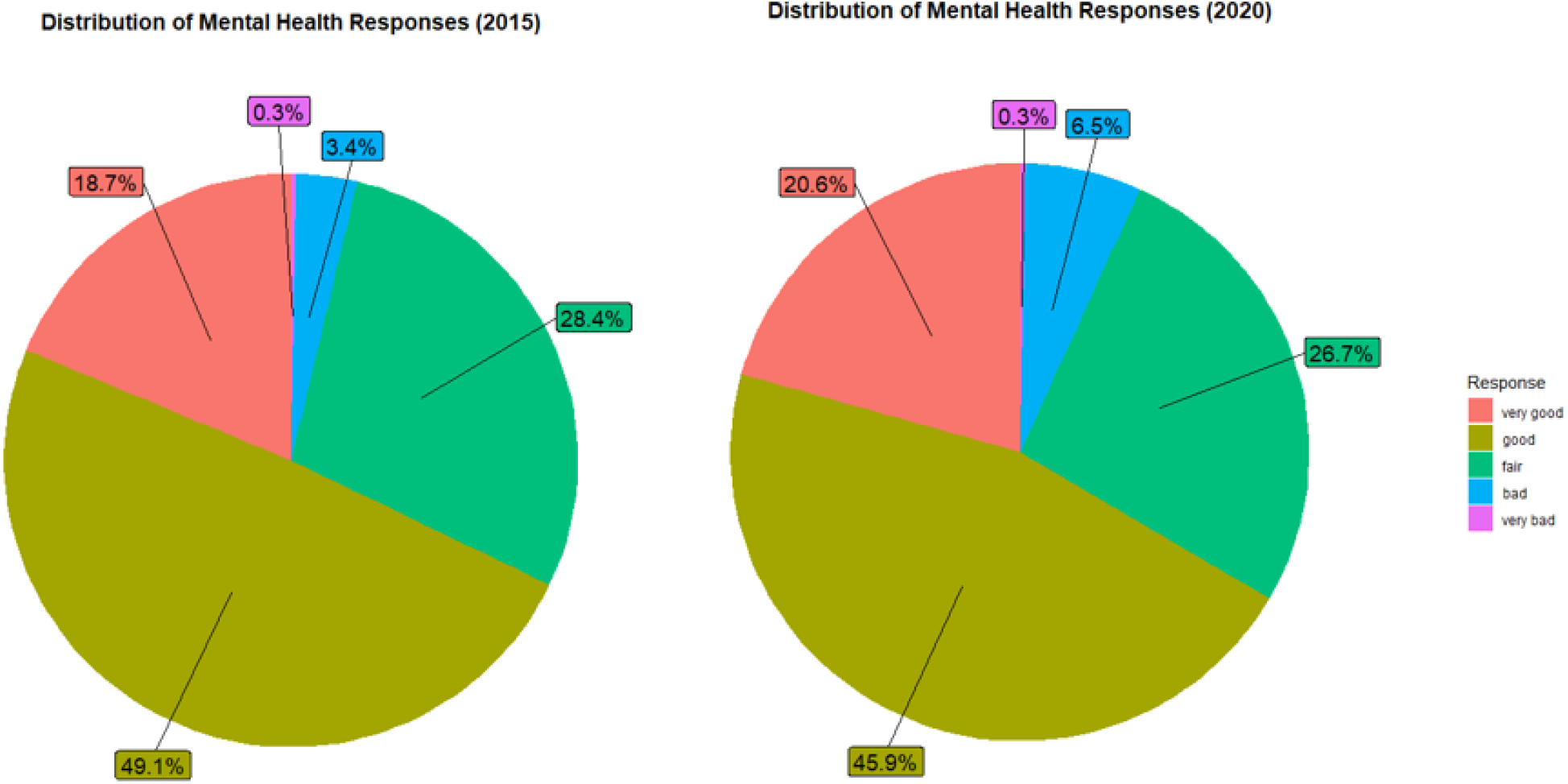
Comparison of distribution of responses for Likert-type mental health item

## Discussion

In general, there was higher self-reported alcohol consumption in 2015 than in 2020, which is in line with previous research on the effect of COVID on alcohol consumption as well as the trends in APC (Kilian, et al., 2022; Kilian, et al., 2021). Although COVID-19 may have had a complex effect on the availability and affordability of alcohol, two key factors in consumption (Babor, et al., 2022), it primarily resulted in reduced consumption. The drop in consumption was not, however, observed ubiquitously across the population. At the highest decile, the trend of higher consumption pre-pandemic was eliminated, suggesting that the heaviest drinkers saw a significant increase in consumption during COVID. When this effect was separated by sex, at the highest decile, females consumed as much alcohol as males during 2020, which was not the case in 2015. These findings support the polarization hypothesis, and suggest that the gap between the sexes for average daily consumption among the heaviest consumers closed during the pandemic.

Although the mechanism of why this occurs is unclear, the present study hints at some possible reasons. Firstly, females appeared to dramatically increase their consumption in 2020. Assuming this trend is generalizable to other countries, the increase in consumption in females is a novel finding to note, since males typically consume more than females (Moinuddin et al., 2016). The analysis of the self-reported mental health item also suggests that mental health could have had an influence in the increased consumption as poor mental health is correlated to increased alcohol consumption (Jacob et al., 2021). A more thorough investigation of the mechanism of why the highest deciles increased alcohol consumption would broaden our understanding of these findings.

Overall, despite the dramatic changes as a result of COVID-19, there remained a significant burden of alcohol consumption on the health of heavy drinkers. Importantly, the high level of consumption was seen in a non-negligible segment of the population (10% of the total population). Still, there are some limitations of the present study. Although many characteristics of Lithuania lend itself to being a comparable example to other EU and Western countries, it is unique in that there is a high overall level of alcohol consumption. Thus, the upper decile of the population, on average, may have a higher level of consumption than in other countries. Second, the analysis was conducted on a general survey that involved two waves of different participants. Thus, although we infer that consumption increased amongst the upper deciles, it is merely speculative (stronger support would come from a direct measure of consumption for the same individuals pre-and post-pandemic). Finally, as with all self-reported measures, there is some margin of error with respect to response biases (e.g., social desirability bias). Thus converging evidence from direct measures of consumption in heavy drinkers from other sources (e.g., clinicians, purchase data, etc.) rather than self-report alone, would strengthen the polarization hypothesis.

COVID was associated with dramatic changes to factors affecting alcohol consumption— from a public policy perspective and from a mental health perspective. Although on a whole, there was a decrease in alcohol consumption, certain segments of the population were affected adversely. We demonstrate, using Lithuania as an example, that vulnerable populations (heavy drinkers, at the upper decile of consumption) had significant increases in consumption which is in line with hypotheses of previous researchers. This finding emphasizes the need to monitor and protect those prone to high alcohol consumption during times of crisis.

## Data Availability

All data produced in the present study are available upon reasonable request to the authors

## Acknowledgements

The authors would like to acknowledge the National Institute on Alcohol Abuse and Alcoholism (NIAAA) (Award Number 1R01AA028224) of the National Institutes of Health for funding this research.

## Ethics Statement

All analyses received Centre for Addiction and Mental Health (CAMH) Research Ethics Board (REB) approval as per protocol: #050/2020.

## Conflict of interest statement

All authors declare no competing interests.

## Author Statement

AT led the conception and design of the study, extracted relevant variables from the dataset, conducted the statistical analyses and contributed to the writing of the manuscript. HJ, JR, SL assisted with the statistical analysis, interpretation of results, and revised the intellectual content of the manuscript. MS, DS, and IT provided the original dataset, produced syntax for the variables, translated the data and revised the intellectual content of the manuscript. All authors approved the final version of this manuscript. AT and DS and IT can verify the underlying data.

## Notes

### Competing Interest Statement

The authors have declared no competing interest.

### Author Declarations

Ethics Statement All analyses received Centre for Addiction and Mental Health (CAMH) Research Ethics Board (REB) approval as per protocol: #050/2020.

## References

Babor, T. F., Casswell, S., Graham, K., Huckle, T., Livingston, M., Österberg, E., … Sornpaisarn, B. (2022). Alcohol: no ordinary commodity: research and public policy.

Bloomfield, K., Kilian, C., Manthey, J., Rehm, J., Brummer, J., & Grittner, U. (2022). Changes in alcohol use in Denmark during the initial months of the COVID-19 pandemic: further evidence of polarization of drinking responses. European Addiction Research, 28(4), 297–308.

Coelho, C. M., Suttiwan, P., Arato, N., & Zsido, A. N. (2020). On the nature of fear and anxiety triggered by COVID-19. Frontiers in psychology, 11, 581314.

De Goeij, M. C., Suhrcke, M., Toffolutti, V., van de Mheen, D., Schoenmakers, T. M., & Kunst, A. E. (2015). How economic crises affect alcohol consumption and alcohol-related health problems: a realist systematic review. Social science & medicine, 131, 131–146.

Erbiçer, E. S., Metin, A., Çetinkaya, A., & şen, S. (2022). The relationship between fear of COVID-19 and depression, anxiety, and stress. European Psychologist.

Ganesan, B., Al-Jumaily, A., Fong, K. N., Prasad, P., Meena, S. K., & Tong, R. K.-Y. (2021). Impact of coronavirus disease 2019 (COVID-19) outbreak quarantine, isolation, and lockdown policies on mental health and suicide. Frontiers in psychiatry, 12, 565190.

Hettmansperger, T. P., & McKean, J. W. (2010). Robust nonparametric statistical methods: CRC press.

Jacob, L., Smith, L., Armstrong, N. C., Yakkundi, A., Barnett, Y., Butler, L., … Meyer, J. (2021). Alcohol use and mental health during COVID-19 lockdown: A cross-sectional study in a sample of UK adults. Drug and alcohol dependence, 219, 108488.

Kilian, C., O’Donnell, A., Potapova, N., López-Pelayo, H., Schulte, B., Miquel, L., … Rehm, J. (2022). Changes in alcohol use during the COVID-19 pandemic in Europe: A meta-analysis of observational studies. Drug and alcohol review, 41(4), 918–931.

Kilian, C., Rehm, J., Allebeck, P., Braddick, F., Gual, A., Barták, M., … O’Donnell, A. (2021). Alcohol consumption during the COVID-19 pandemic in Europe: a large-scale cross-sectional study in 21 countries. Addiction, 116(12), 3369–3380.

Lilliefors, H. W. (1967). On the Kolmogorov-Smirnov test for normality with mean and variance unknown. Journal of the American statistical Association, 62(318), 399–402.

McBride, E., Arden, M. A., Chater, A., & Chilcot, J. (2021). The impact of COVID-19 on health behaviour, well-being, and long-term physical health. British Journal of Health Psychology, 26(2), 259.

Moinuddin, A., Goel, A., Saini, S., Bajpai, A., & Misra, R. (2016). Alcohol consumption and gender: a critical review. J Psychol Psychother, 6(3), 1–4.

Neufeld, M., Lachenmeier, D. W., Ferreira-Borges, C., & Rehm, J. (2020). Is alcohol an “Essential Good” during COVID-19? Yes, but only as a disinfectant! Alcoholism, Clinical and Experimental Research, 44(9), 1906.

Onyeaka, H., Anumudu, C. K., Al-Sharify, Z. T., Egele-Godswill, E., & Mbaegbu, P. (2021). COVID-19 pandemic: A review of the global lockdown and its far-reaching effects. Science progress, 104(2), 00368504211019854.

Pohorecky, L. A. (1991). Stress and alcohol interaction: an update of human research. Alcoholism: Clinical and experimental research, 15(3), 438–459.

R Development Core Team. (2023). R: A language and environment for statistical computing (Version 4.3.1). Vienna, Austria: R Foundation for Statistical Computing. Retrieved from https://www.R-project.org/

Rehm, J. (2024). Covid-19, the polarization of substance use, and mental health. Jornal de Pediatria, 100(4).

Rehm, J., Kilian, C., Ferreira-Borges, C., Jernigan, D., Monteiro, M., Parry, C. D., … Manthey, J. (2020). Alcohol use in times of the COVID 19: Implications for monitoring and policy. Drug and alcohol review, 39(4), 301–304.

Sohi, I., Chrystoja, B. R., Rehm, J., Wells, S., Monteiro, M., Ali, S., & Shield, K. D. (2022). Changes in alcohol use during the COVID-19 pandemic and previous pandemics: A systematic review. Alcoholism: clinical and experimental research, 46(4), 498–513.

Verma, B. K., Verma, M., Verma, V. K., Abdullah, R. B., Nath, D. C., Khan, H. T., … Verma, V. (2020). Global lockdown: An effective safeguard in responding to the threat of COVID-19. Journal of evaluation in clinical practice, 26(6), 1592–1598.

World Health Organization. (2024). Global status report on alcohol and health and treatment of substance use disorders. Retrieved from Geneva, Switzerland:

